# Clinical Trials for Wolfram Syndrome Neurodegeneration: Novel Design, Endpoints, and Analysis Models

**DOI:** 10.1101/2024.09.10.24313426

**Authors:** Guoqiao Wang, Zhaolong Adrian Li, Ling Chen, Heather Lugar, Tamara Hershey

## Abstract

**Objective:** Wolfram syndrome, an ultra-rare condition, currently lacks effective treatment options. The rarity of this disease presents significant challenges in conducting clinical trials, particularly in achieving sufficient statistical power (e.g., 80%). The objective of this study is to propose a novel clinical trial design based on real-world data to reduce the sample size required for conducting clinical trials for Wolfram syndrome.

**Methods:** We propose a novel clinical trial design with three key features aimed at reducing sample size and improve efficiency: (i) Pooling historical/external controls from a longitudinal observational study conducted by the Washington University Wolfram Research Clinic. (ii) Utilizing run-in data to estimate model parameters. (iii) Simultaneously tracking treatment effects in two endpoints using a multivariate proportional linear mixed effects model.

**Results:** Comprehensive simulations were conducted based on real-world data obtained through the Wolfram syndrome longitudinal observational study. Our simulations demonstrate that this proposed design can substantially reduce sample size requirements. Specifically, with a bivariate endpoint and the inclusion of run-in data, a sample size of approximately 30 per group can achieve over 80% power, assuming the placebo progression rate remains consistent during both the run-in and randomized periods. In cases where the placebo progression rate varies, the sample size increases to approximately 50 per group.

**Conclusions:** For rare diseases like Wolfram syndrome, leveraging existing resources such as historical/external controls and run-in data, along with evaluating comprehensive treatment effects using bivariate/multivariate endpoints, can significantly expedite the development of new drugs.

## 1. Introduction

Wolfram syndrome is a rare (estimated to affect 1 in 770,000 to 160,000 people), autosomal recessive, multisystem disease first defined in 1938 as the combination of childhood-onset insulin-dependent diabetes, optic nerve atrophy, diabetes insipidus, and hearing loss [1, 2]. The major causative gene *WFS1* encodes for wolframin [3], an endoplasmic reticulum (ER) transmembrane glycoprotein involved in preventing cell death potentially through suppressing ER stress [4], regulating intracellular calcium homeostasis [5], and modulating mitochondrial function [6]. It is now understood that pathogenic mutations in *WFS1* can result in death or dysfunction of insulin-producing pancreatic β cells [3], causing insulin-dependent diabetes. *WFS1* mutations are also presumed to impact other organ systems [7], leading to a wide range of symptomatology. Several agents proposed to protect cells from ER stress-mediated apoptosis are being tested [8-13]; however, currently there are no approved treatments that alter Wolfram syndrome progression.

The identification of causative *WFS1* genetic mutations has allowed for a genetic rather than purely clinical diagnosis of Wolfram syndrome. Observations of genetically diagnosed Wolfram syndrome individuals indicate that the disease phenotype is indeed much more variable than classically understood, extending to loss of taste and smell [14-16], bladder and bowel dysfunction [17-19], gait and balance issues [20, 21], and mental health disorders [14, 22]. Wolfram syndrome is associated with early brain structural alterations such as reduced or stalled white matter development [23-26]. In addition, patterns of explicit neurodegeneration can be detected in gray matter regions over 2-3 years, regardless of age [24]. As these abnormalities progress and brain-mediated symptoms worsen, quality of life decreases and life-threatening conditions can develop (e.g., respiratory failure from brainstem atrophy) [27].

Identifying biomarkers of brain degeneration is thus a critical step towards designing efficient and effective clinical trials for candidate pharmaceutical agents or therapies. We have conducted the world’s largest and longest natural history study of early Wolfram syndrome, focusing on quantification of neurological features of the disease and their changes over time. Over the 13 years of the Washington University Wolfram Research Clinic (https://wolframsyndrome.wustl.edu/items/research-clinic/; ClinicalTrials.gov Identifier NCT03951298), we validated a standardized clinical severity rating scale for Wolfram syndrome (the Wolfram Unified Rating Scale, WURS) [28], described unexpectedly early neurological symptoms and neuropathological differences, and characterized the rate of change in these symptoms and in regional brain volumes. We found that visual acuity and the volume of the thalamus (based on magnetic resonance imaging [MRI]) deteriorate the most consistently and rapidly in our cohort [25, 29]. This information has been used to justify outcome measures and predict power for safety[8] and clinical efficacy studies using standard designs (e.g., ClinicalTrials.gov Identifier: NCT03717909).

One of the primary challenges for clinical trials in rare diseases such as Wolfram syndrome is enrolling an adequate number of participants to ensure sufficient statistical power. Here, we leverage our unique longitudinal natural history dataset to explore the added power obtained (and reduced sample sizes required) by using three different innovative design features. First, we consider the impact of including historical data from patients who were not treated in the clinical trial. This approach allows more participants to receive active treatment without compromising statistical power compared to the traditional 1:1 randomization trial [30, 31]. For example, a trial with 2:1 randomization can be analyzed as if it had a 1:1 ratio by borrowing the same number of historical/external controls as the randomized placebo controls. Second, we consider how best to use data collected from Wolfram syndrome patients prior to their participation in a clinical trial (i.e., ‘run-in’ data [32, 33]). Run-in data aids in estimating important model parameters such as the rate of change of the placebo group or the variances, thereby enhancing statistical power [32, 33]. Depending on whether the disease progression remains relatively stable during a moderate follow-up duration (e.g., a total of 4-6 years), the disease progression rate for participants on placebo can be assumed to be the same or different during the run-in period and the randomized period (**Figure 1**).

**Figure 1:**
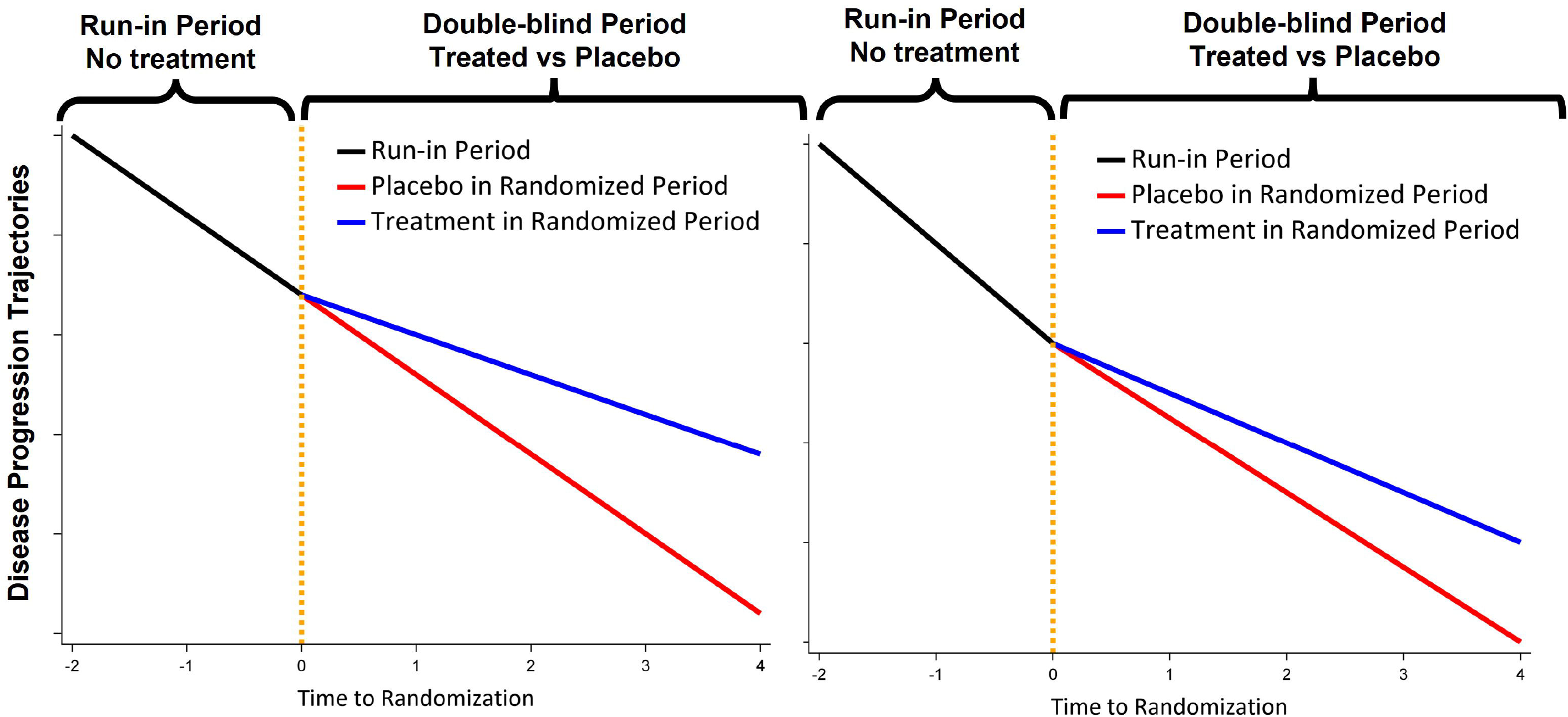
Demonstration of how clinical trials might use run-in data. Left: a trial design that uses the same disease progression rate for placebo during the run-in period and the randomized period; Right: a trial design that uses different disease progression rates for placebo during the run-in period and the randomized period.

Finally, a single primary endpoint may not fully capture the important effects of an intervention, particularly for diseases with diverse manifestations such as Wolfram syndrome. In such cases, the use of multiple primary endpoints presents an appealing solution [34]. The multiple endpoints can be modeled in several ways: as a multivariate endpoint [35], as a composite endpoint (e.g., average [31, 36] or sum [37]), or by analyzing each endpoint individually in a sequential manner. The use of an integrated scale such as a composite endpoint [38, 39] has occurred in Alzheimer’s disease trials and can be acceptable to the FDA [40]. However, a composite endpoint may impose challenges in interpretation and may inadvertently assign disproportionate weight to a particular endpoint upon averaging or summing [41] . Analyzing multiple endpoints sequentially requires careful control of type I error and can sometimes result in a loss of statistical power [42]. To address these limitations, we propose a direct modeling approach that considers multiple endpoints simultaneously using proportional mixed models for repeated measures [43]. This approach allows for the identification of a single proportional reduction in disease progression across multiple endpoints, effectively capturing the treatment effect across a range of measures.

In this paper, we present the impact of these three design features on the predicted power and required sample sizes in clinical trials of interventions that target neurodegeneration in Wolfram syndrome. By using our unique longitudinal dataset from a natural history study of Wolfram syndrome, we can empirically determine the design that optimally balances the power to detect slowing of neurodegeneration while minimizing the required sample size. Future clinical trials targeting neurodegeneration in Wolfram syndrome may then use this information to optimize their trial designs and justify their proposed sample sizes.

## 2 Methods

### 2.1 Data

Participants with genetically confirmed *WFS1* mutations, and under the age of 30 at enrollment, were assessed annually at the Washington University Wolfram Syndrome Research Clinic[24]. Briefly, at each annual visit participants were evaluated by a pediatric ophthalmologist or optometrist who measured best corrected visual acuity by Snellen optotype [29]. In the following analyses, we used the logarithm of the minimum angle of resolution (logMAR) value for best-corrected visual acuity with both eyes open; higher logMAR value indicates worse visual acuity. Participants also underwent magnetic resonance imaging where a T1-weighted high-resolution magnetization-prepared rapid gradient-echo (MPRAGE) sequence was acquired. Scans from each annual visit were processed longitudinally using the semi-automatic segmentation program FreeSurfer v5.3 [44], and regional brain volumes were extracted and corrected for estimated total intracranial volume (eTIV) [45]. Data acquired between 2010 and 2017 were included in these analyses.

### 2.2 Models

#### 2.2.1 Univariate Linear Mixed Effects Model

The univariate linear mixed effects (LME) model that estimates a linear disease progression rate can be written as [43, 46]:

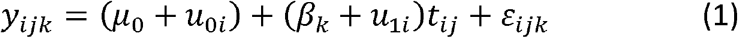

Where *y*_*ijk*_ denotes the longitudinal assessments for subject *i* at time *j* for group *k*,*i*=1,2,…,*n*,*j* =0,1…,*n*_*i*_ with *j*=0 representing the baseline visit, and k=1,2 representing the placebo group and the treatment group, respectively*;μ*_0_ is the baseline intercept,β_*k*_ is the disease progression rate for group *k* where time *t*_*ij*_ represents subject-specific assessment time;u_0*i*_,u_1*i*_are the random effects for the intercept and the progression rate (i.e., slope) and are assumed to follow the same bivariate normal distribution for both groups: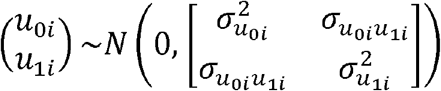 the within-subject error is assumed to follow the same normal distribution for both groups 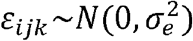.

#### 2.2.2 Treatment Effect Represented by Difference in Disease Progression Rates or Proportional/Percent Reduction

When analyzing the primary endpoint using model (1), the treatment effect is commonly expressed as the difference between the annual disease progression rates, i.e., *β*_1_ − *β*_2_ for the two subject groups. An alternative approach is to model the treatment effect as the proportional/percent reduction in the disease progression rate relative to the placebo rate, as described below [30, 43]:

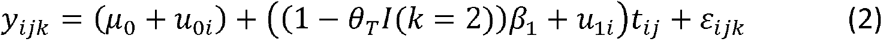

whereθ*T* is the proportional/percentage reduction treatment effect; *I* is the indicator function with *I* (*k*=2)=1 and *I* (*k*=1)=0;*β*_1_ is the disease progression rate of the placebo group, and all the other parameters have the same meanings as those in model (1).

#### 2.2.3 Multivariate Proportional Linear Mixed Effects Model

Utilizing a proportional/percent reduction to represent the treatment effect offers several advantages: 1) enhances the interpretability of the treatment effect compared to solely focusing on the difference in slope of the absolute values; 2) enables the totality evaluation of the overall treatment effect across multiple endpoints; and 3) allows for a reduction in sample size by simultaneously modeling multivariate endpoints. Such multivariate endpoints can be modeled using a multivariate proportional mixed effects model as below, which accounts for the correlation among different endpoints through the use of random effects [43]:

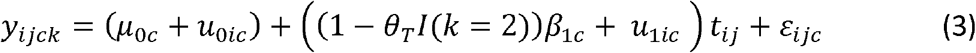

Where, *i* represents the subject; *j* represents the visit/time where *j*=0 at baseline; *c* is the index for the endpoint included in the multivariate endpoint; *k*=1,2 represents the placebo group and the treatment group, respectively; *I* is the indicator function with I(*k*=2)=1and I(*k*=1)=0; μ_0*c*_and*β*_*1C*_represent the baseline intercept and the progression rate of the placebo group for endpoint *c*, respectively;*u*_0*ic*_,*u*_1*ic*_ are the random effects for the intercept and the progression rate (slope) for subject *i* endpoint *c*, and are assumed to follow a multivariate normal distribution N(0_2*C*,_ Σ_*2C×2C*_)(e.g., 4 by 4 unstructured covariance matrix if there are 2 endpoints);θ_*T*_ is the treatment effect represented as a proportional/percentage reduction of the placebo disease progression; and *ε*_*ijc*_ is the within-subject residual and has different variances for different endpoints.

## 3. Results

Characteristics of participants in the Wolfram syndrome longitudinal observational study have been previously described [24]. In the current study, data from a total of 37 patients genetically diagnosed with Wolfram syndrome were included. Among them, 21 individuals (57%) were female. The mean (SD) baseline age of the participants was 13.3 (5.5) years, and the mean (SD) follow-up duration was 2.9 (1.2) years.

### 3.1 Estimated Annual Disease Progression Rates and the Associated Variance/Covariance

A bivariate linear mixed-effects model was employed to estimate the annual progression rates and the associated variance/covariance components for both visual acuity and thalamus volume simultaneously in participants with Wolfram syndrome. These two biomarkers were found to deteriorate the most consistently and rapidly in previous longitudinal observational data [25, 29], suggesting usefulness as disease progression endpoints. To facilitate model convergence, thalamus volumes were normalized to z-scores using the baseline mean (SD) of healthy controls. This adjustment made the range of normalized thalamus volume and visual acuity more compatible. Estimates are presented in **Table 1** and **Table 2**. For interpretability, the estimated intercept for visual acuity of 0.540 logMAR corresponded to about 20/69 ft (6/21 m; 3^rd^ line) on the Snellen scale, and the estimated slope of 0.062 logMAR/y translated to approximately 7.4 years to deteriorate to 20/200 ft (6/60 m; 1^st^ line).

**Table 1:** Estimated annual disease progression rates and 95% CI for each endpoint.

| Parameter | Estimate | SE | P-value | 95% Confidence Limits |  |
| --- | --- | --- | --- | --- | --- |
| Intercept of Visual Acuity (logMAR) | 0.540 | 0.069 | <.0001 | 0.399 | 0.681 |
| Slope of Visual Acuity (LogMAR/year) | 0.062 | 0.013 | <.0001 | 0.035 | 0.089 |
| Intercept of Thalamus Volume (z-score) | -1.116 | 0.165 | <.0001 | -1.451 | -0.781 |
| Slope of Thalamus Volume (z-score/year) | -0.139 | 0.023 | <.0001 | -0.185 | -0.093 |

**Table 2:** Estimated variance/covariance components for each endpoint.

|  | Intercept of Visual Acuity | Slope of Visual Acuity | Intercept of Thalamus Volume | Slope of Thalamus Volume |
| --- | --- | --- | --- | --- |
| Intercept of Visual Acuity | 0.174 | 0.020 | -0.178 | -0.020 |
| Slope of Visual Acuity | 0.020 | 0.004 | -0.042 | -0.002 |
| Intercept of Thalamus Volume | -0.178 | -0.042 | 0.811 | 0.025 |
| Slope of Thalamus Volume | -0.020 | -0.002 | 0.025 | 0.007 |
| Residual Variance of Visual Acuity | 0.025 |  |  |  |
| Residual Variance of Thalamus Volume | 0.005 |  |  |  |

### 3.2 Evaluation of Model Performance by Study Designs and Endpoints

To evaluate model performance for each study design (i.e., historical controls, run-in data) and each type of endpoints (i.e., univariate, multivariate), we conducted simulations for various scenarios. The simulations were configured as follows:

- Individual baseline values and annual progression rates for visual acuity and thalamus volume were generated using a multivariate normal distribution with the mean and variance/covariance components specified in **Table 1** and **Table 2**.
- Individual longitudinal data were simulated by combining the individual baseline values,annual progression rates, and within-subject error, according to model 3.
- Treatment effects were introduced as a proportional reduction in the placebo rate of progression at various levels: 0% (Type I error), 30%, and 40%.
- Time was set to 0 at baseline, negative before baseline (i.e., the run-in period), and positive after baseline.
- The run-in period lasted for 2 years, and the treatment period lasted for 3 years.
- Assessments were scheduled every 6 months.
- The annual dropout rate was set at 10%.
- The randomization ratio was 2:1 with pooled historical/external controls to make the power estimation based on a randomization ratio of 1:1, the sample sizes range from 30 per group to 120 per group.
- For each model, 1000 data sets were simulated, allowing for estimation of power with precision up to three decimal places [47].
- Type I error and power were calculated as the proportion of the 1000 simulated trials per scenario that yielded P-values < 0.05.
- Disease progression rates during both the run-in and randomized periods are the same vs different (slower progression during the randomized period due to the placebo effect).

#### 3.2.1 Type I Error/Power Estimation with vs. without Run-in Data for a Single Primary Endpoint (Visual Acuity)

Figure 2. illustrates the power of various design and analysis models. The inclusion of run-in data significantly enhances power compared to trials without run-in data, regardless of whether the placebo progression rates remain the same or not throughout both run-in and randomized periods. Nevertheless, when the placebo progression rates are the same during run-in and randomized periods (**Figure 2A** and **2B**), utilization of run-in data results in a greater increase in power compared to scenarios where the placebo progression rates change (**Figure 2C** and **2D**). Estimating the treatment effect as a percent reduction demonstrates higher power compared to estimating it as the difference in disease progression rates. Moreover, such gain in power is more pronounced when the disease progression rates for placebo participants differ between the run-in and randomized periods. The type I error for all designs/models generally remains within 2% of the nominal 5% level (Supplemental Figure 1).

**Figure 2:**
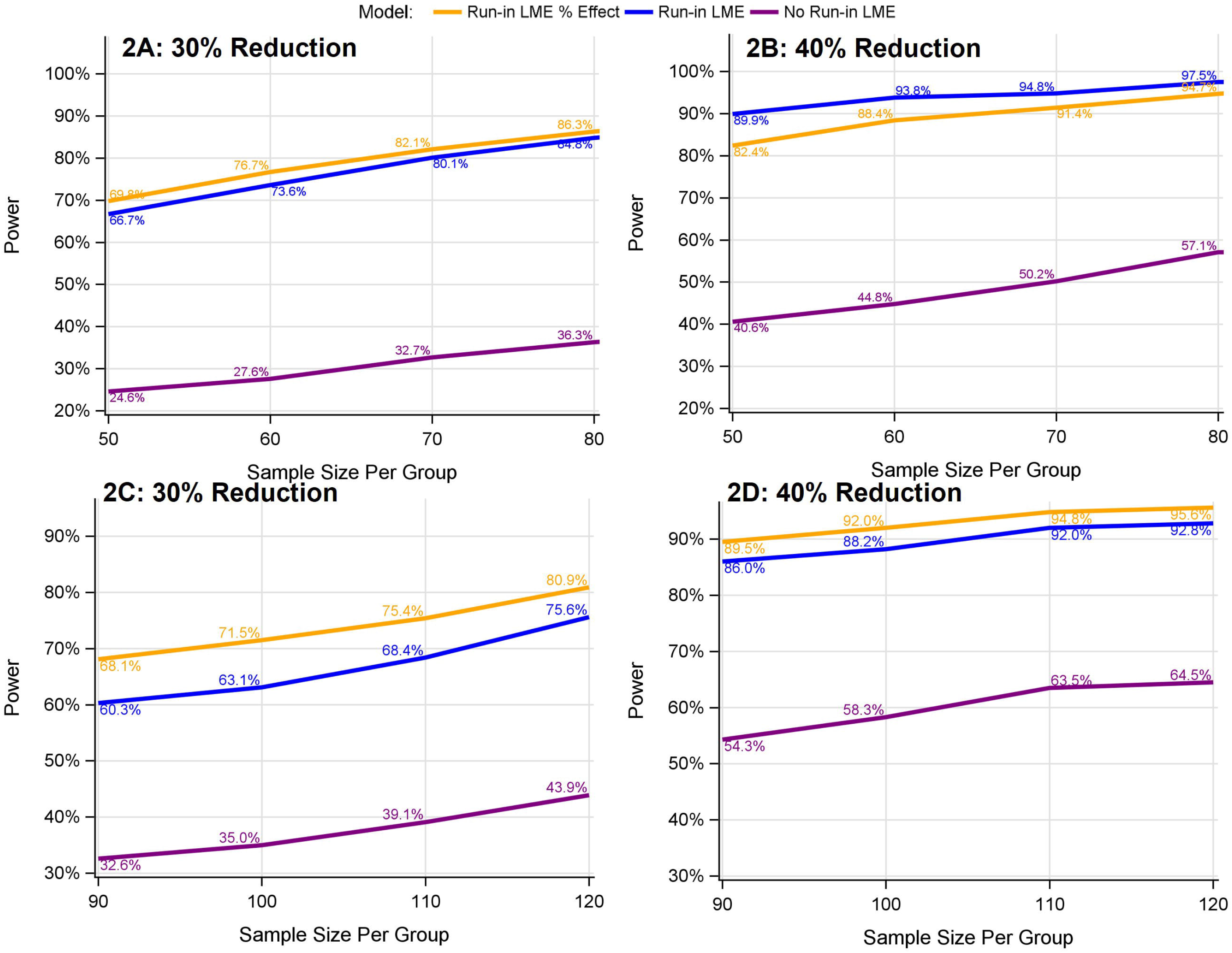
Power comparison between models with vs. without run-in data for the univariate endpoint of visual acuity. **A** and **B:** The disease progression rate for placebo participants is assumed to be the same throughout both the run-in and randomized periods (as in left panel in **Figure 1**). **C** and **D**: The disease progression rate for placebo participants is assumed to be slower in the randomized than the run-in period (as in right panel in **Figure 1**). 30%, 40% reduction: 30%, 40% reduction in the disease progression relative to the placebo group. No Run-in LME (purple lines): without run-in data analyzed using LME model 1. Run-in LME (blue lines): with run-in data analyzed using LME model 1. Run-in LME % Effect: (yellow lines) with run-in data analyzed using LME model 2

#### 3.2.2 Type I Error/Power Estimation for Bivariate Endpoint (Visual Acuity and Thalamus Volume)

Figure 3. presents an illustration of power associated with bivariate endpoints, with or without run-in data. Consistent with the simulations conducted for the univariate endpoint models, the inclusion of run-in data enhances power considerably. Further, the bivariate endpoint provides a significant gain in power, effectively doubling or even tripling the power observed with the univariate endpoint. Overall, the type I error for all endpoints/designs typically stays within 2% of the expected 5% level (Supplemental Figure 1).

**Figure 3:**
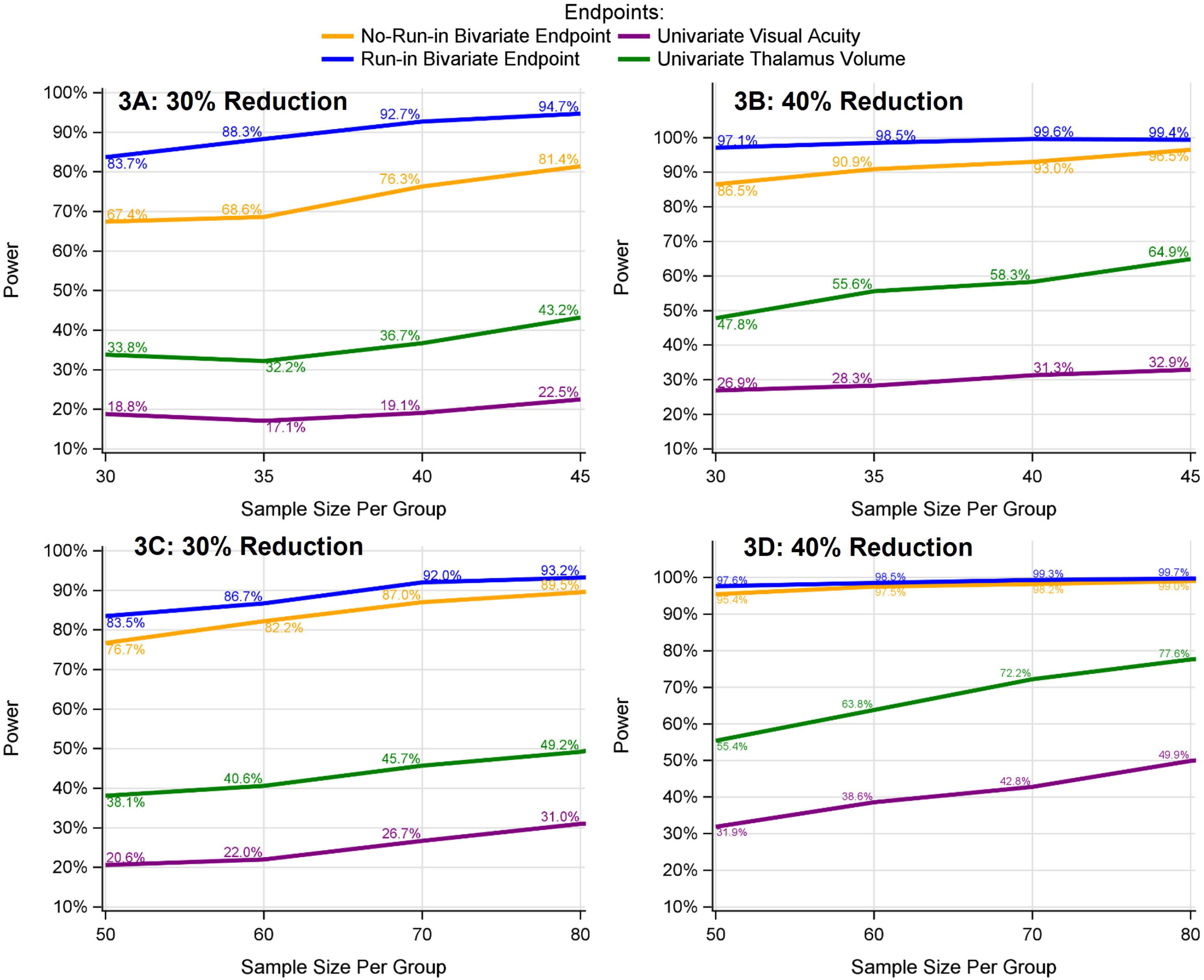
Power comparison between models with bivariate vs. univariate endpoints and with vs. without run-in data. **A** and **B**: The disease progression rate for placebo participants is assumed to be the same throughout both run-in and randomized periods (as in left panel in **Figure 1). C** and **D**: The disease progression rate for placebo participants is assumed to be slower during the randomized than run-in period (as in right panel in **Figure 1**). 30%, 40% reduction: 30%, 40% reduction in the disease progression relative to the placebo group. Univariate Visual Acuity: visual acuity analyzed by LME model 1. Univariate Thalamus Volume: thalamus volume analyzed by LME model 1; No-Run-in Bivariate Endpoint: bivariate endpoint of visual acuity and thalamus volume analyzed by model 3 without run-in data; Run-in Bivariate Endpoint: bivariate endpoint of visual acuity and thalamus volume analyzed by model 3 with run-in data.

## 4. Discussion

Our results demonstrate the range of power that can be obtained in clinical trials targeting neurodegeneration in Wolfram syndrome using practical and data-driven approaches. By leveraging our unique existing natural history dataset, we demonstrate the considerable potential gain in power achieved by using run-in and historical/external control data and by choosing a bivariate primary endpoint, assuming a 3-year follow-up with 6-month assessment intervals. After taking into account the assumptions and caveats associated with these models, investigators will be able to use this information and maximize existing resources to guide and standardize clinical trial protocols going forward, leading to more efficient trial designs for this ultra-rare disease.

The three features of the proposed design that contributed to increased power were the borrowing of historical/external controls, use of run-in data, and a bivariate primary endpoint of visual acuity and thalamus volume. Each of these design features boosted our power estimates, although there are some additional issues that need to be discussed. First, by leveraging historical data from our natural history study, we can employ an unequal randomization ratio, such as 2:1 with two times the number of patients on the active treatment. Our simulations assumed that we could borrow enough historical/external controls so that the efficacy analysis would have a 1:1 randomization ratio. In a real-world trial, however, the availability of historical/external controls will vary. Second, although each individual had 2 years of run-in data in the simulation, it is not necessary that the run-in data have equal duration. Any amount of run-in data with various durations can help [32]. In any trial that includes run-in data, it is recommended to conduct tailored simulations to evaluate the specific power gain from these run-in data.

Finally, our simulations indicated that relying on a single endpoint is not ideal for such a rare disease with a wide spectrum of symptomatology. A bivariate endpoint (visual acuity and thalamus volume) not only significantly improved statistical power, but also would provide a more comprehensive evaluation of treatment effects across the disease spectrum without compromising type I error control. However, the bivariate endpoint method assumes that the treatment effect is approximately the same across endpoints. Although this assumption might seem strong, it has been implicitly used in various composite endpoints [30, 31, 48]. When the proportional treatment effect differs between endpoints, the model will estimate an average treatment effect across all endpoints, similar to the use of a composite score of averaging multiple endpoints [30, 31, 48]. We recommend conducting a sensitivity bivariate analysis to estimate the proportional treatment effect for each endpoint separately. By comparing the average treatment effect to endpoint-specific treatment effects, a comprehensive evaluation of the treatment effect can be achieved.

While not directly addressed in our simulations, it may be beneficial for the field to consider a platform trial with a master protocol for early Wolfram syndrome. A platform trial is a method for evaluating multiple targeted therapies within a single disease setting, using a perpetual framework where therapies can enter or exit the platform based on predefined decision algorithms [49]. Building platform trials with master protocols for Wolfram syndrome will enable more efficient utilization of resources and accelerate the pace of drug discovery. Such trials facilitate the establishment of a trial network infrastructure that encourages extensive collaboration among researchers in the field of rare diseases [49]. Currently, a group of investigators across Europe are conducting a multi-site clinical trial (ClinicalTrials.gov Identifier:

NCT03717909) with a master protocol. We would advocate for a similar approach within the US, ideally calibrating measures across sites, clinics and regions.

Our study has several limitations. First, our natural history data were obtained from a non-random group of participants. Enrolled participants were generally young, in an early disease state, and able to travel to St. Louis for annual several day-long visits. In addition, they were able to perform visual acuity tests (i.e., were not already blind) and did not have any contraindications for MRI scans (e.g., no cochlear implants, bladder stimulators, etc.). As a result, older, more impaired participants were not included in the study. However, the younger and less impaired individuals are the ones that presumably would benefit the most from slowing neurodegeneration and avoiding irreversible consequences of Wolfram syndrome. Lastly, there may be other, unexplored measures of neurodegeneration that could be more precise or more easily collected than MRI-based regional brain volumes. For example, fluid biomarkers are becoming more available and acceptable in clinical trials for other neurodegenerative conditions [38, 50] and may have promise in Wolfram syndrome [51].

In this paper, we propose a novel paradigm for conducting clinical trials in Wolfram syndrome, focusing on optimizing available resources and statistical approaches to enhance the feasibility of trials in this rare disease. The use of historical/external controls, run-in data, and a bivariate endpoint can greatly reduce the sample size required, thus facilitating the progress towards identifying truly effective drugs for this rare disease.

## Data Availability

The raw data used in the analyses described in this manuscript cannot be made available in the manuscript, supplemental files, or a public repository due to the small sample size and rarity of Wolfram syndrome, which could lead to the identification of individuals even after data de-identification. The corresponding author may be contacted to request data. According to the Human Research Protection Office (HRPO) at Washington University, a preface to a data sharing agreement and a data sharing agreement reviewed by the research office will be employed prior to data sharing. HRPO regulations permit access to potentially identifiable data only to research personnel on our study protocol and approved through the University.

## List of Abbreviations

ER: endoplasmic reticulum
WURS: Wolfram Unified Rating Scale
MRI: magnetic resonance imaging
logMAR: logarithm of the minimum angle of resolution
eTIV: estimated total intracranial volume
LME: linear mixed effects.

## Declarations

## Ethics approval and consent to participate

Not applicable

## Consent for publication

All authors consent for publication

## Authors’ contributions

All authors (GW, ZL, LC, HL, and TH) contributed to the conception, writing, and editing of this manuscript. GW conducted all the simulations.

## Acknowledgements

None

## References

1. Dj, W. Diabetes mellitus and simple optic atrophy among siblings: Report of 4 cases. in Mayo Clin Proc. 1938.

2. Barrett, T.G., S.E. Bundey, and A.F. Macleod, Neurodegeneration and diabetes: UK nationwide study of Wolfram (DIDMOAD) syndrome. The Lancet, 1995. 346(8988): p. 1458–1463.

3. Inoue, H., et al., A gene encoding a transmembrane protein is mutated in patients with diabetes mellitus and optic atrophy (Wolfram syndrome). Nature genetics, 1998. 20(2): p. 143–148.

4. Shang, L., et al., beta-cell dysfunction due to increased ER stress in a stem cell model of Wolfram syndrome. Diabetes, 2014. 63(3): p. 923–33.

5. Lu, S., et al., A calcium-dependent protease as a potential therapeutic target for Wolfram syndrome. Proceedings of the National Academy of Sciences, 2014. 111(49): p. E5292–E5301.

6. Zmyslowska, A., et al., Multiomic analysis on human cell model of wolfram syndrome reveals changes in mitochondrial morphology and function. Cell Commun Signal, 2021. 19(1): p. 116.

7. Urano, F., Wolfram syndrome: diagnosis, management, and treatment. Current diabetes reports, 2016. 16: p. 1–8.

8. Abreu, D., et al., A phase Ib/IIa clinical trial of dantrolene sodium in patients with Wolfram syndrome. JCI Insight, 2021. 6(15).

9. Abreu, D. and F. Urano, Current Landscape of Treatments for Wolfram Syndrome. Trends Pharmacol Sci, 2019. 40(10): p. 711–714.

10. Gorgogietas, V., et al., GLP-1R agonists demonstrate potential to treat Wolfram syndrome in human preclinical models. Diabetologia, 2023. 66(7): p. 1306–1321.

11. Lu, S., et al., A calcium-dependent protease as a potential therapeutic target for Wolfram syndrome. Proc. Natl. Acad. Sci. U. S. A, 2014. 111(49): p. E5292–E5301.

12. Mahadevan, J., et al., A soluble endoplasmic reticulum factor as regenerative therapy for Wolfram syndrome. Lab Invest, 2020. 100(9): p. 1197–1207.

13. Nguyen, L.D., et al., Calpain inhibitor and ibudilast rescue beta cell functions in a cellular model of Wolfram syndrome. Proc Natl Acad Sci U S A, 2020. 117(29): p. 17389–17398.

14. Bischoff, A.N., et al., Selective cognitive and psychiatric manifestations in Wolfram Syndrome. Orphanet. J Rare. Dis, 2015. 10(1): p. 66.

15. Alfaro, R., et al., Taste and smell function in Wolfram syndrome. Orphanet J Rare Dis, 2020. 15(1): p. 57.

16. Alfaro, R., et al., Enhancement of taste by retronasal odors in patients with Wolfram syndrome and decreased olfactory function. Chem Senses, 2023. 48.

17. Doty, T., et al., The effects of disease-related symptoms on daily function in Wolfram Syndrome. Transl Sci Rare Dis, 2017. 2(1-2): p. 89–100.

18. Marshall, B.A., et al., Phenotypic characteristics of early Wolfram syndrome. Orphanet. J Rare. Dis, 2013. 8: p. 64.

19. Rove, K.O., et al., Lower Urinary Tract Dysfunction and Associated Pons Volume in Patients with Wolfram Syndrome. J Urol, 2018.

20. Pickett, K.A., et al., Early presentation of gait impairment in Wolfram Syndrome. Orphanet. J Rare. Dis, 2012. 7: p. 92.

21. Pickett, K.A., et al., Balance impairment in individuals with Wolfram syndrome. Gait. Posture, 2012. 36(3): p. 619–624.

22. Reiersen, A.M., et al., Psychiatric Diagnoses and Medications in Wolfram Syndrome. Scand J Child Adolesc Psychiatr Psychol, 2022. 10(1): p. 163–174.

23. Hershey, T., et al., Early brain vulnerability in Wolfram syndrome. PLoS. One, 2012. 7(7): p. e40604.

24. Lugar, H.M., et al., Evidence for altered neurodevelopment and neurodegeneration in Wolfram syndrome using longitudinal morphometry. Sci Rep, 2019. 9(1): p. 6010.

25. Lugar, H.M., et al., Neuroimaging evidence of deficient axon myelination in Wolfram syndrome. Scientific reports, 2016. 6(1): p. 21167.

26. Samara, A., et al., Developmental hypomyelination in Wolfram syndrome: new insights from neuroimaging and gene expression analyses. Orphanet J Rare Dis, 2019. 14(1): p. 279.

27. Minton, J.A.L., et al., Wolfram syndrome. Reviews in Endocrine & Metabolic Disorders, 2003. 4(1): p. 53–59.

28. Nguyen, C., et al., Reliability and validity of the Wolfram Unified Rating Scale (WURS). Orphanet. J Rare. Dis, 2012. 7: p. 89.

29. O’bryhim, B.E., et al., Longitudinal Changes in Vision and Retinal Morphology in Wolfram Syndrome. American journal of ophthalmology, 2022. 243: p. 10–18.

30. Wang, G., et al., A novel cognitive disease progression model for clinical trials in autosomal-dominant Alzheimer’s disease. Statistics in medicine, 2018.

31. Bateman, R.J., et al., The DIAN-TU Next Generation Alzheimer’s prevention trial: Adaptive design and disease progression model. Alzheimer’s & Dementia, 2017. 13(1): p. 8–19.

32. Wang, G., et al., Two-period linear mixed effects models to analyze clinical trials with run-in data when the primary outcome is continuous: Applications to Alzheimer’s disease. Alzheimer’s & Dementia: Translational Research & Clinical Interventions, 2019. 5: p. 450–457.

33. Madsen, K., J. Miller, and M. Province, The use of an extended baseline period in the evaluation of treatment in a longitudinal Duchenne muscular dystrophy trial. Statistics in medicine, 1986. 5(3): p. 231–241.

34. Hamasaki, T., S.R. Evans, and K. Asakura, Design, data monitoring, and analysis of clinical trials with co-primary endpoints: a review. Journal of biopharmaceutical statistics, 2018. 28(1): p. 28–51.

35. Salloway, S., et al., A trial of gantenerumab or solanezumab in dominantly inherited Alzheimer’s disease. Nature medicine, 2021. 27(7): p. 1187–1196.

36. Papp, K.V., et al., Sensitivity of the preclinical Alzheimer’s cognitive composite (PACC), PACC5, and repeatable battery for neuropsychological status (RBANS) to amyloid status in preclinical Alzheimer’s disease-atabecestat phase 2b/3 EARLY clinical trial. The Journal of Prevention of Alzheimer’s Disease, 2022. 9(2): p. 255–261.

37. Mintun, M.A., et al., Donanemab in early Alzheimer’s disease. New England Journal of Medicine, 2021. 384(18): p. 1691–1704.

38. Salloway, S., et al., A trial of gantenerumab or solanezumab in dominantly inherited Alzheimer’s disease. Nature Medicine, 2021: p. 1–10.

39. Sims, J.R., et al., Donanemab in early symptomatic Alzheimer disease: the TRAILBLAZER-ALZ 2 randomized clinical trial. JAMA, 2023.

40. Morant, A.V., et al., US, EU, and Japanese regulatory guidelines for development of drugs for treatment of Alzheimer’s disease: implications for global drug development. Clinical and Translational Science, 2020. 13(4): p. 652–664.

41. Xiong, C., et al., Combining multiple markers to improve the longitudinal rate of progression: application to clinical trials on the early stage of Alzheimer’s disease. Statistics in Biopharmaceutical Research, 2013. 5(1): p. 54–66.

42. Guidance, D., Multiple endpoints in clinical trials guidance for industry. Center for Biologics Evaluation and Research (CBER), 2017.

43. Wang, G., et al., Proportional constrained longitudinal data analysis models for clinical trials in sporadic Alzheimer’s disease. Alzheimer’s & Dementia: Translational Research & Clinical Interventions, 2022. 8(1): p. e12286.

44. Fischl, B., et al., Whole brain segmentation: automated labeling of neuroanatomical structures in the human brain. Neuron, 2002. 33(3): p. 341–355.

45. Buckner, R.L., et al., A unified approach for morphometric and functional data analysis in young, old, and demented adults using automated atlas-based head size normalization: reliability and validation against manual measurement of total intracranial volume. Neuroimage, 2004. 23(2): p. 724–38.

46. Fitzmaurice, G.M., N.M. Laird, and J.H. Ware, Applied longitudinal analysis. Vol. 998. 2012: John Wiley & Sons.

47. Wang, G., et al., Effect of sample size re-estimation in adaptive clinical trials for Alzheimer’s disease and mild cognitive impairment. Alzheimer’s & Dementia: Translational Research & Clinical Interventions, 2015. 1(1): p. 63–71.

48. Van Dyck, C.H., et al., Lecanemab in early Alzheimer’s disease. New England Journal of Medicine, 2023. 388(1): p. 9–21.

49. Woodcock, J. and L.M. LaVange, Master protocols to study multiple therapies, multiple diseases, or both. New England Journal of Medicine, 2017. 377(1): p. 62–70.

50. Wang, G., et al., Staging biomarkers in preclinical autosomal dominant Alzheimer’s disease by estimated years to symptom onset. Alzheimer’s & Dementia, 2019. 15(4): p. 506–514.

51. Eisenstein, S.A., et al., Plasma Neurofilament Light Chain Levels Are Elevated in Children and Young Adults With Wolfram Syndrome. Front Neurosci, 2022. 16: p. 795317.

