## Supplemental Figure 1 for "Clinical Trials for Wolfram Syndrome Neurodegeneration: Novel Design, Endpoints, and Analysis Models"

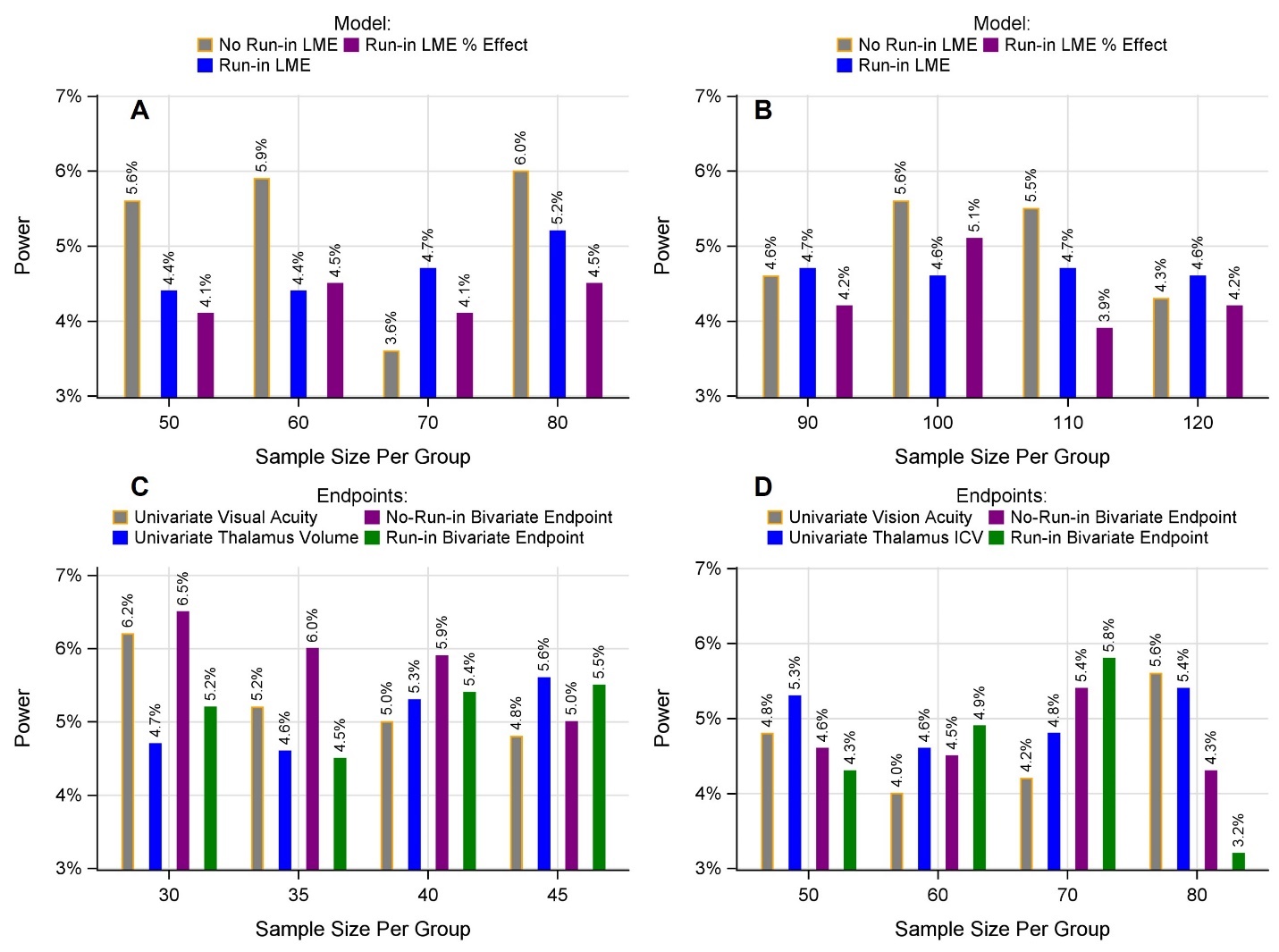

**Supplemental Figure 1**: Type I error by sample size for different endpoints/models. A: Single primary endpoint, placebo progression rates remain the same during both the run-in and randomized periods; B: Single primary endpoint, placebo progression rates are different during the run-in and randomized periods; C: Placebo progression rates remain the same during both the run-in and randomized periods; D: Placebo progression rates are different during the run-in and randomized periods.
